# Beyond geography: a catchment-based typology of rural and coastal urgent and emergency care sites in England

**DOI:** 10.64898/2026.10.27.26366554

**Authors:** Wei Song, Joy Adamson, James Gaughan, Peter Lampard, Daniel Mensah, Gerry Richardson, Arabella Scantlebury, Peter Sivey, Rita Santos, the ELEVATE team

## Abstract

Reducing geographical inequalities in urgent and emergency care requires identifying services that support rural and coastal populations. Classifying services by hospital location may misrepresent the communities they serve. We developed a catchment-based typology based on the area characteristics of the population that emergency services serve, rather than their postcode.

Using NHS England Emergency Care Data Set for April 2021-March 2024, we linked individual attendances to the patient’s residential area rurality and coastal proximity. Site-level rural and coastal attendance shares were compared with national averages using single-proportion tests. We then assessed variations in service pressures across location classes by analysing attendance volume, seasonality, and patient characteristics.

The study included 63,924,015 attendances across 347 sites, of which 344 were assigned to four classes: rural-coastal, rural-inland, urban-coastal and urban-inland. Rural-coastal sites had lower average attendance volumes with pronounced summer increases, driven by a higher proportion of attendances from patients living in urban and inland areas. Urban sites (coastal and inland) had higher average proportions of ambulance arrivals and high-acuity attendances. Comparisons of site-level and attendance-weighted measures highlighted heterogeneity within rural classes, with distinct compound demand pressures in rural and coastal UEC services across the year, both in terms of timing and case-mix. The typology provides a transparent basis for service planning, monitoring geographical inequalities and evaluating whether policy and service innovations have different effects across service contexts.

## 1 Background

Urgent and Emergency Care (UEC) services act as both a healthcare safety net of last resort and a crucial barometer of broader health system stress. While mitigating geographical disparities in access and outcomes, especially for rural and coastal populations, is a central objective across OECD countries, health systems lack a practical mechanism to identify which UEC facilities predominantly serve these communities. This gap limits our ability to evaluate place-based health policies.

Demand for UEC services varies between places and over time, reflecting different population health needs, service accessibility, and how urgent care capacity is distributed between UEC services and alternative routes into care. Rural and coastal communities experience poorer access to care, driven by lower service density and extended travel times relative to urban centres (Xiao-dong et al., 2022; Mseke et al., 2024). They are also more likely to have older populations and more complex health and care needs. Coastal communities are frequently associated with poorer underlying health and wider health inequalities (Asthana and Gibson, 2022). Tourism and temporary residency generate predictable intra-year surges in healthcare demand, even where the baseline resident population remains stable. In England, these seasonal peaks are highly concentrated in coastal regions during the summer.(Turcato et al., 2023).

Despite these longstanding concerns, routine monitoring and much of the empirical literature still classify healthcare services primarily by their postcode rather than by the population they serve. This distinction matters because, while emergency care is delivered at specific provider sites, facility locations often misalign with catchment area characteristics. An inland hospital may draw heavily from coastal populations, whereas a coastal site may treat substantial numbers of inland residents or temporary visitors. A UEC site that is geographically inland may serve a substantial coastal population, while a coastal site treat substantial numbers of inland towns and cities residents or temporary visitors.As a result, headline performance metrics can conceal contrasting operational realities. This measurement gap is especially pronounced for rurality and coastal proximity, metrics established for small geographical areas but rarely mapped to health service sites (MacKinney, A. Clinton et al., 2014). Existing classifications, including those based on official small-area definitions, are therefore not well suited to identify which UEC sites actually serve rural and/or coastal populations, or to monitor and evaluate policy at the site level (Welsh et al., 2024).

This paper addresses this gap by developing a catchment-based typology of UEC sites in England, defined by the residential geography of attending patients rather than provider location. Using small-area administrative data for all attendances, we construct site-level catchment profiles and evaluate each facility’s rural and coastal attendance shares against national benchmarks. We produce four classes: Rural & Coastal, Rural & Inland, Urban & Coastal, and Urban & Inland. The approach aligns with a larger literature on defining catchment areas using observed patient flows, which has been used in other settings to delineate hospital service areas and referral regions (Meakin and Funk, 2024; Jacob et al., 2024; Santos et al., 2020).

The value of a catchment-based typology is threefold. First, it provides a transparent way to identify which UEC sites serve urban/rural and/or inland/coastal communities, defined by who a site serves rather than where it sits. Second, it becomes feasible to test whether seasonal pressures, case-mix, waiting-time performance, or other system performance indicators differ systematically between urban/rural and inland/coastal UEC sites. Third, it allows researchers and policymakers to assess whether the impact of policies intended to improve UEC performance and reduce geographical disparities has different effects across services serving urban/rural and inland/coastal communities (Lampard et al., 2026). In doing so, the typology can support more policy-relevant comparisons across England, allowing better evaluation of service innovation and recovery initiatives and more informed decision-making on resource allocation. This will provide a foundation for future work on the effectiveness and cost-effectiveness of innovations in UEC services that serve rural and coastal communities.

## 2 Methods

We conduct a national observational study of urgent and emergency care (UEC) attendances in England to classify UEC sites according to the rurality and coastalness of the populations they serve. The unit of analysis is the UEC site, defined as the emergency care service recorded in the Emergency Care Data Set (ECDS) and listed on NHS England’s Organisation Data Service (ODS) portal, including the postcode (NHS England Digital, 2026). Patient residence is identified using Lower Super Output Area (LSOA) of residence, a small-area geography as part of the national census in England. These areas are defined by the Office for National Statistics (ONS) for the Census in 2001, 2011 and 2021. Each LSOA has an average population of around 1,500 people spread around 400-1,200 households at the time of the Census.

### 2.1 Catchment profiling

We characterise each UEC site by mapping patient attendances to the rural and coastal classifications of their residential Lower Layer Super Output Area (LSOA). Aggregating these data, we calculate the proportion of total attendances at each UEC originating from rural and coastal LSOAs. We then compare each site’s rural-attendance share and coastal-attendance share with the corresponding national attendance shares over the study period using one-sided single-proportion tests (Kirkwood, Betty R. and Sterne, Jonathan A. C., 2003). Sites are assigned to one of four classes (Rural & Coastal, Rural & Inland, Urban & Coastal, Urban & Inland) depending on whether their rural and/or coastal attendance shares are statistically higher than the national average. Details are included in Appendix B. A small number of sites (3 sites, accounting for 1.28% of attendances) are not assigned to any class because their rural and/or coastal attendance shares are not statistically different from the national averages. These sites are excluded from class-based analyses.

### 2.2 Data sources and linkage

Our main source of data is the Emergency Care Data Set (ECDS), provided by NHS England. We analyse all attendances to UEC in England from April 2021 to March 2024. ECDS is an attendance-level dataset that captures patient characteristics, the NHS trust site attended, timestamps along the emergency care pathway, diagnoses, investigations, and treatments administered during the patient’s attendance. The data includes each patient’s LSOA of residence.

In order to define rurality and coastalness, we link ECDS to two datasets at the LSOA level (version 2021) using each patient’s residential LSOA. First, we link the ONS 2021 Rural-Urban Classification to identify the rurality of patients’ LSOAs of residence (Office for National Statistics, 2026b). ONS developed a Rural-Urban Classification for English and Welsh LSOAs based on settlement size and sparsity (Office for National Statistics, 2026a). Of the six ONS categories, we consider LSOAs within the four rural categories (RLN1, RSN1, RSF1, and RLF1). Second, we link the NHS Digital dataset on straight-line distances from population-weighted LSOA centroids to the nearest coastline to define coastal LSOAs (NHS Digital, 2024). After mapping these distances to LSOA 2021, we define an LSOA as coastal if its centroid lies within 5 kilometres (km) of the coastline. Our choice of a 5 km threshold is consistent with recent work in England showing that coastal definitions based on a 5 km shoreline buffer best distinguish communities with distinct health profiles, including cancer incidence and prevalence (Welsh et al., 2024). This threshold also reflects a pragmatic consideration that coastal populations are often served by sites located slightly inland rather than directly on the coastline. An LSOA is considered rural and coastal if it meets both criteria.

We also link ECDS to the Income Deprivation Score and Rank at LSOA 2011 from the English Indices of Deprivation (Ministry of Housing, Communities and Local Government, 2019). LSOA 2011 is used as deprivation scores were released before the 2021 census. We use the Organisational Data Service (ODS) data to identify provider mergers and service reconfigurations over the study period. To ensure consistency over time, we harmonise provider and site identifiers to a stable definition based on the earliest period of observation, so organisational changes do not create artificial variation in the set of UEC sites ^1^.

In addition, some urgent and emergency care services are co-located at the same physical site (e.g. emergency departments and urgent treatment centres) but may be recorded as separate site identifiers in ECDS depending on local reporting practices. Because these services often operate within a single physical location and reporting practices have changed over time, we harmonise co-located services to a single site identifier at postcode level. This ensures that differences in reporting do not affect the definition of a site ^2^.

## 3 Results

We identified 421 UEC provider sites across the three financial years 2021/22 to 2023/24 of ECDS. After accounting for UEC provider mergers and reorganisations during the study period, postcode site harmonisation, and removal of UEC sites with an average of less than 100 attendances per year, there are 347 provider UEC sites. These account for over 63,924,000 attendances over the three financial years. Across all sites, 14.58% of attendances are from a rural LSOA, and 16.44% are from a coastal LSOA.

Table 1 summarises the four classes identified using our catchment-based typology.

**Table 1:** Class-level descriptive statistics (2021/22–2023/24).

|  | Rural &<br>Coastal | Rural &<br>Inland | Urban &<br>Coastal | Urban &<br>Inland |
| --- | --- | --- | --- | --- |
| Sites | 67 | 121 | 42 | 114 |
| Sites reporting Type 1 activity: n (% of sites) | 26 (38.8%) | 62 (51.2%) | 25 (59.5%) | 77 (67.5%) |
| Site-years | 196 | 356 | 121 | 333 |
| Total annual attendances | 2,608,185 | 5,990,073 | 2,745,283 | 9,692,059 |
| Annual Type 1 attendances: n (%) | 1,816,796 (69.7%) | 4,613,329 (77.0%) | 1,951,837 (71.1%) | 7,470,231 (77.1%) |
| Annual attendances per site: mean (SD) | 38,981 (35,223) | 49,642 (43,392) | 66,315 (47,805) | 85,331 (51,939) |
| Annual attendances per site: median [IQR] | 26,485 [9,035–60,880] | 28,236 [12,521–83,379] | 62,757 [21,001–111,537] | 90,582 [43,395–127,684] |
| Rural attendances (%) | 36.5 | 35.9 | 6.1 | 3.6 |
| Coastal attendances (%) | 61.2 | 1.6 | 68.6 | 1.2 |

SD: standard deviation. IQR: interquartile range. Type 1 site counts include any site reporting Type 1 activity during the study period, including sites also reporting other department types. Type 1 attendance counts include only records labelled Type 1; annual counts are three-year totals divided by three. Percentages use the corresponding class totals.

For each class, we report the number of sites and observed site-years, defined as one site observed in one financial year, the total annual number of attendances, and the typical site size. These include the mean annual attendances per site and the median number of annual attendances, with interquartile ranges. The final columns give the share of attendance from patients living in rural and coastal LSOAs. 67 out of the 347 sites (19.3%) are classed as Rural & Coastal, given the significantly higher attendance from patients living in rural and coastal areas. The distribution is skewed toward urban & inland in both number of sites and per-site volume of attendances. Rural & coastal comprise fewer and smaller sites.

The type of UEC services also differed across classes. The proportion of sites reporting Type 1 activity ranged from 38.8% in Rural Coastal to 67.5% in Urban & Inland. Nevertheless, Type 1 activity accounted for most attendances in every class, ranging from 69.7% to 77.1%.

Figure 1 shows all 347 sites on a map of England along with their respective classes. From this figure, UEC sites in England are concentrated in urban areas and transport corridors. Large metropolitan areas like Greater London, West Midlands, and North West Yorkshire urban areas have many UEC sites in close geographical proximity. More sparsely populated areas have longer inter-site distances, most notably around the Lake District, the Pennines, and parts of the South West Peninsula. This spatial pattern is consistent with the idea that access to emergency care is partly shaped by geography and population density. In some regions, the nearest site may be substantially further away.

**Figure 1:**
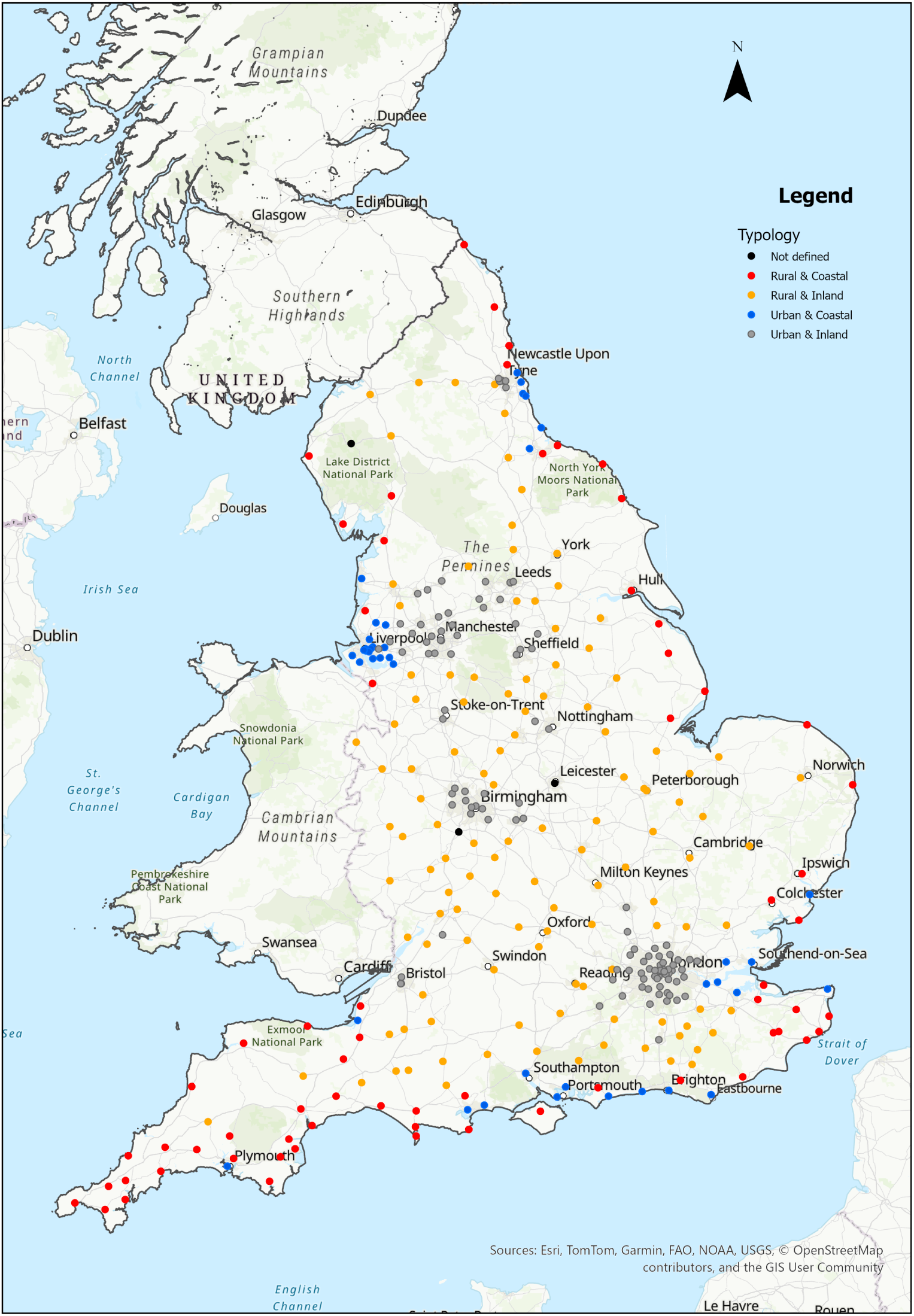
Map of emergency care sites by catchment classes (Rural & Coastal, Rural & Inland, Urban & Coastal, Urban & Inland).

The map shows that Rural & Coastal UEC sites, shown in red, form a near-continuous chain around the South West and much of the southern coastline, particularly visible along Cornwall and Devon and across the south coast. There are additional clusters of Rural & Coastal UEC sites around the North East coast, namely the Yorkshire and Humber and the East Anglia and Kent coastlines. Urban & Inland UEC sites, shown in grey, create the largest cluster in London, with secondary clusters around Birmingham, Manchester, Liverpool, Sheffield and Leeds. There are fewer Urban & Coastal UEC sites, shown in blue which are on or near major coastal urban centres, most visibly along the South Coast, Merseyside and the North East coast. Rural & Inland UEC sites, shown in orange, dominate much of the interior, appearing broadly dispersed across the Midlands and large parts of the East and South East outside the main urban clusters.

### 3.1 Seasonality

Figure 2 summarises within year seasonality in attendances over the 36-month study period. Each site is normalised to its own financial year mean: the index is equal to 1 when the number of attendances in the month equals the site’s annual average. It is above or below 1 when the month is busier or quieter. The series are plotted averages of this index across the UEC sites.

**Figure 2:**
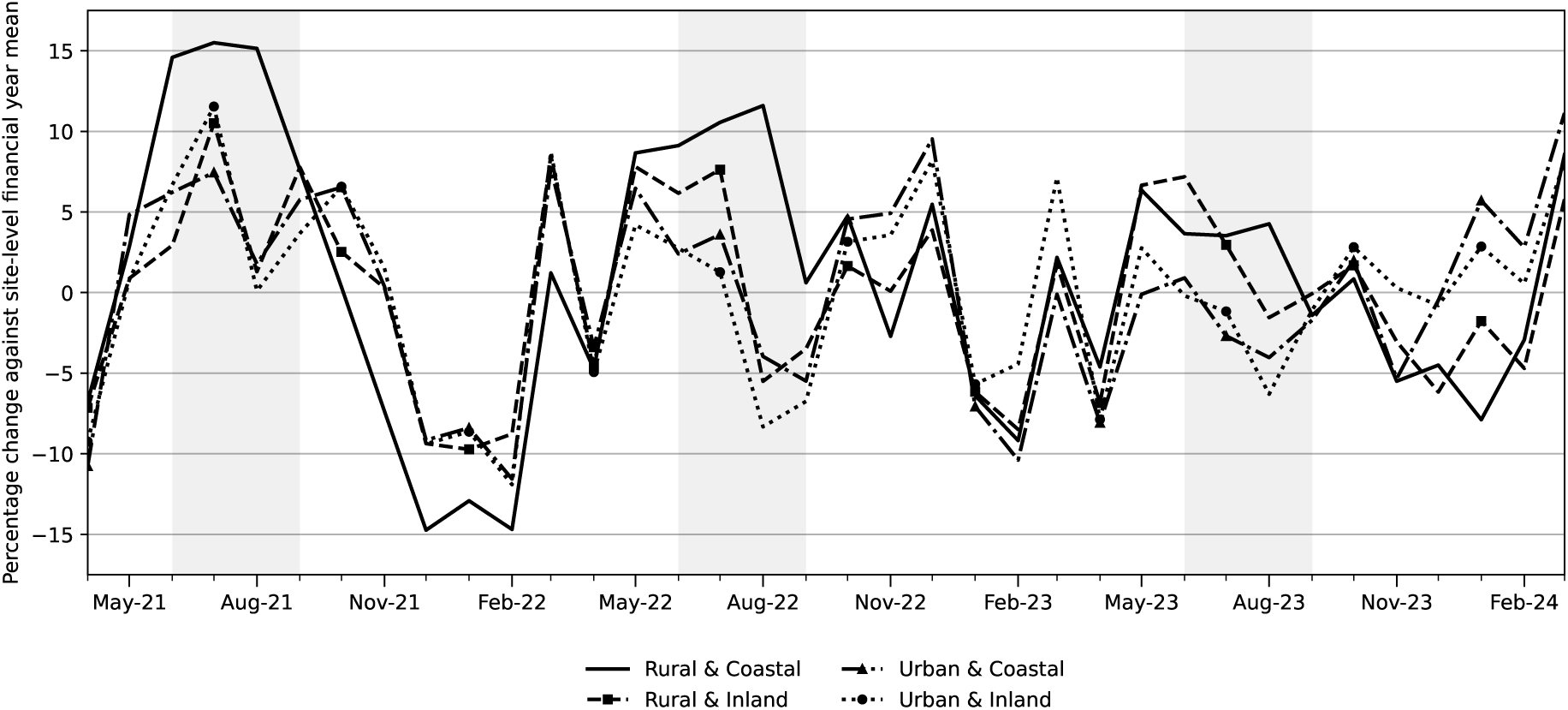
Monthly changes in emergency care utilisation by class, 2021/22 to 2023/24.

The lines allow for a comparison of relative seasonal pressure across the four classes. The shaded bands highlight the summer months (June-August) in each year. Across these shaded months, Rural & Coastal UECs have, on average, 9.2% more attendances than their site-year mean. Rural & Inland UECs have, on average, 3.5% more attendances. Rural & Coastal UECs have the highest number of attendances in July 2021 (+15.5%) and August 2022 (+11.6%). In winter 2021/22, attendance decreased across all classifications, associated with the COVID-19 pandemic and the omicron wave of infection. In the two winters of 2022/23 and 2023/24, there is no noticeable winter decrease, except for Rural & Coastal UEC sites. For these sites, attendances in February are around 14.7% below the mean in 2022 and 20.0% below in 2023. Overall, the pattern for Rural & Coastal UEC sites is consistent with a specific concentration of attendances in the summer period, with fewer attendances in winter.

Figure 3 reports the average monthly attendances per site for each class. Monthly counts are first averaged within a site over all years and then across UEC sites within class. Absolute volumes are highest for Urban & Inland UECs (7,200 attendances per month per site, rounded to the nearest hundred), followed by Urban & Coastal (5,600), Rural & Inland (4,200), and Rural & Coastal (3,300). Within Rural & Coastal UECs, mean monthly volume rises from about 3,000 attendances per site in February to around 3,500 in June, an increase of 483 attendances per site (15.9%).

**Figure 3:**
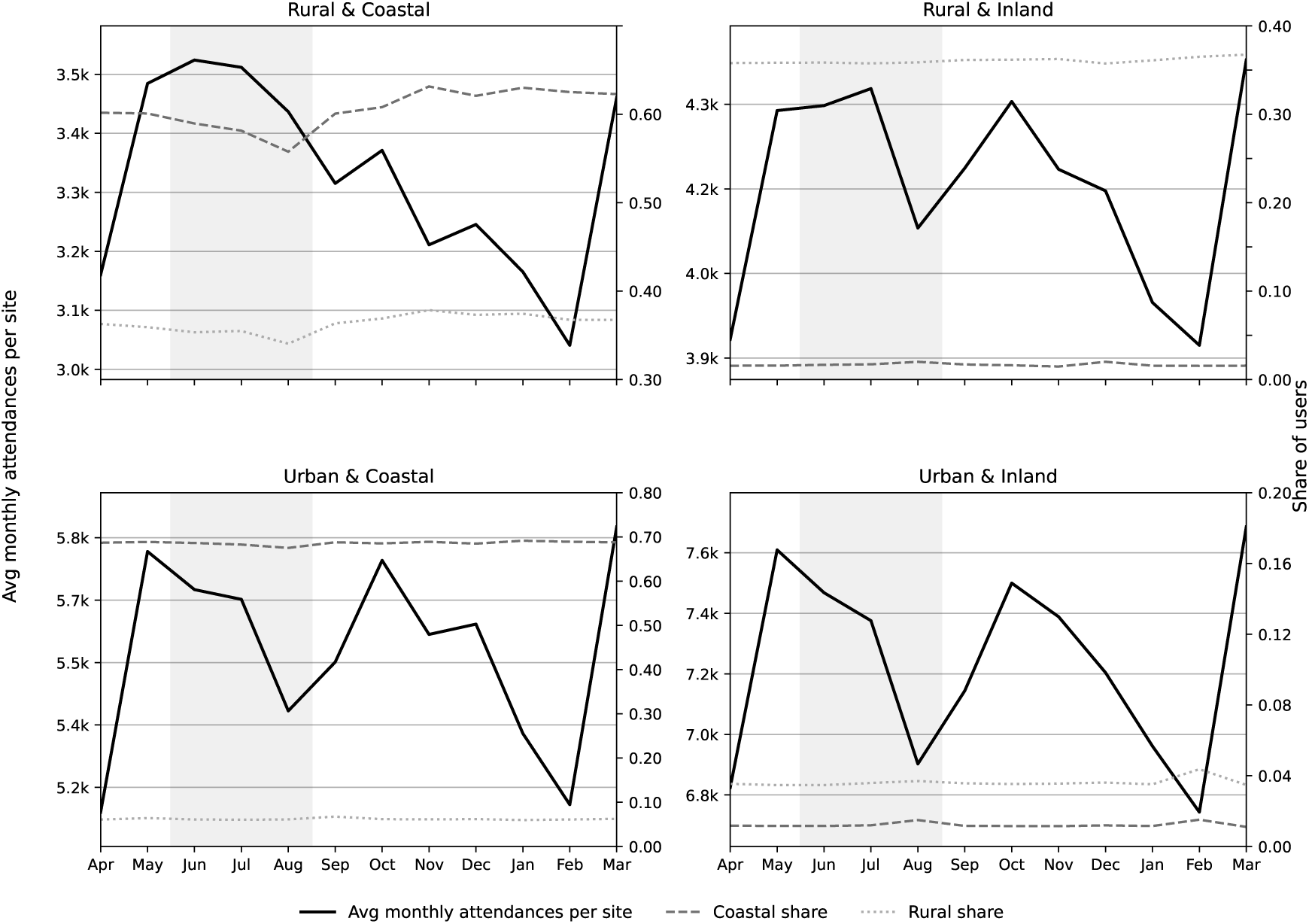
Average monthly attendances per site and coastal/rural user shares.

The right-hand axis overlays the monthly share of attendances from rural and coastal LSOAs.

Shifts indicate a changing mix of who attends. In the Rural & Coastal panel, the increase in volume coincides with decreases in both the rural and coastal shares. This is consistent with additional summer activity being disproportionately generated by users from non-rural and non-coastal LSOAs. This finding contrasts with other classes, where equivalent LSOA type shares are similar over the year. This suggests that seasonal fluctuations for these UECs largely reflect changes in overall demand rather than large shifts in the population attending UEC. Taken together with Figure 2, our findings indicate that Rural & Coastal UECs face a specific challenge in the summer months, partly driven by inflows from patients who are not usual catchment area residents.

### 3.2 Attendance composition and casemix

To describe attendance composition, we use two ECDS variables that proxy patient acuity (arrival by ambulance and high acuity), patient age, and patients’ small area of residence income deprivation. We focus on the percentage of arrivals by road ambulance from the three main available sources (ambulance, own or public transport, and other or unknown) and on the percentage of high-acuity attendances, defined as those with triage levels 1 to 3 from 1 to 5. For each metric, we aggregate to the class level using site means and attendance-weighted means.

Figure 4 shows the attendance composition as the site mean and the attendance-weighted mean for our typology of UEC sites, by attendance arrival mode and acuity, patient age and patient’s small area of residence income deprivation.

**Figure 4:**
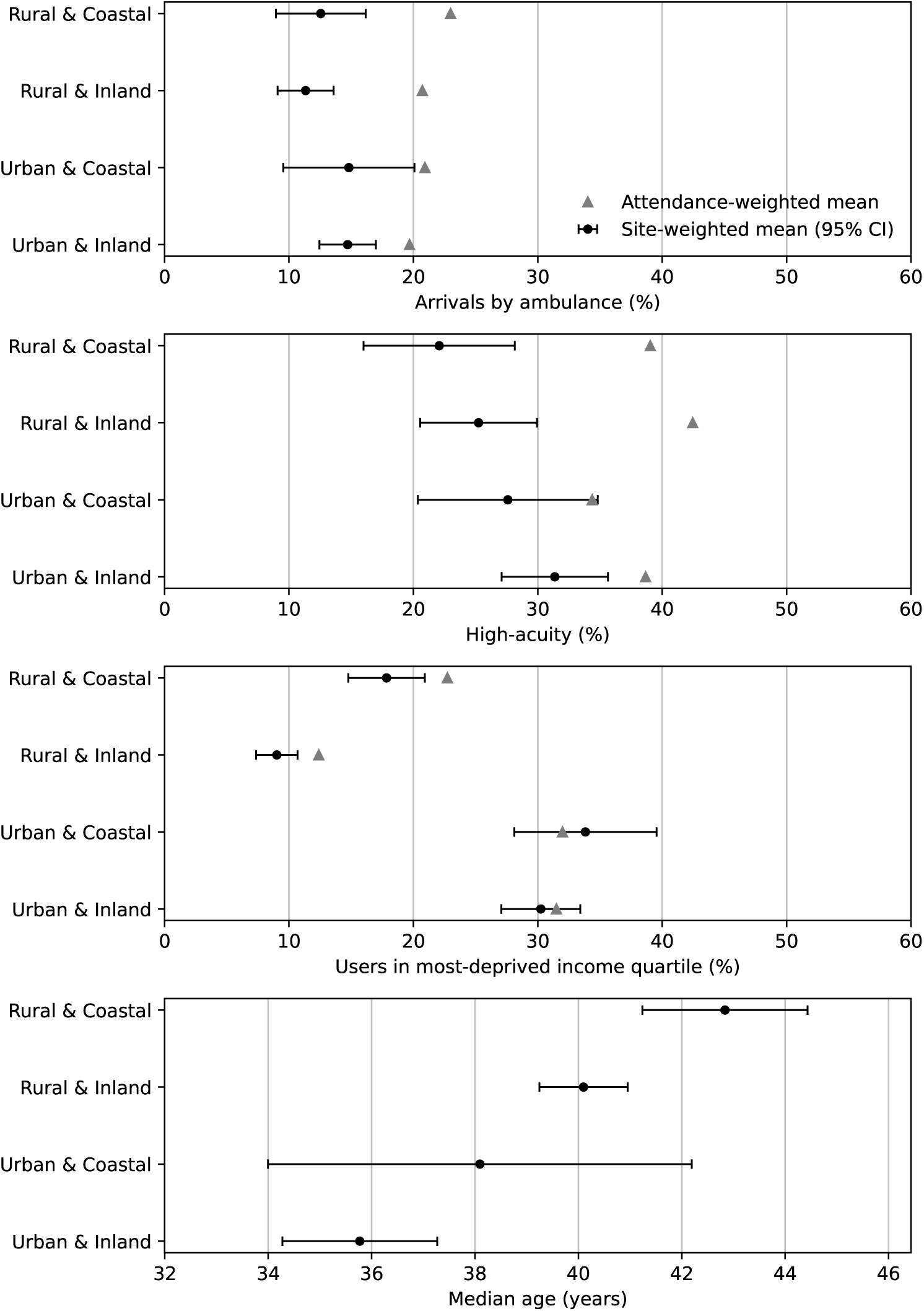
Attendance composition by catchment class: arrivals by ambulance (%), high acuity (%), most-deprived income quartile (%), and median age (years).

In this figure, UEC site means of attendance are notated with circles. These means across UEC describe a ‘typical site’ within the class, with each site weighted equally, and the 95% confidence intervals indicate between-site variation.

Attendance-weighted means are shown in triangles. These describe the composition of attendances, weighting sites by their attendance volume. For a given site class, larger differences between site means and attendance weighted mean suggest the presence of heterogeneity for the specific metric due to the size of UECs within a class. These are more pronounced for arrivals by ambulance and acuity, particularly for the rural-coastal class, as sites of this class that handles a much larger volume of activity compared to a typical rural-coastal site can face a different composition of attendances, with higher proportion of arrivals by ambulance, as well as high acuity.

When comparing across the classes in our typology, circles show differences between typical providers across classes, and triangles show average attendance-weighted differences. Arrival by ambulance is higher in urban UECs (15%) than in rural ones (11% to 12%) on average per UEC site, consistent with the concentration of UECs in urban catchments. For each class, triangles lie to the right of circles, indicating that higher-volume sites have a higher share of ambulance arrivals than lower-volume sites within the same class.

This means that once the volume of the site compared to the typical UEC is taken into account, the Rural & Coastal UEC sites have the highest percentage of arrivals by ambulance. High-acuity attendances show a similar pattern to ambulance arrivals, with urban sites having higher site shares. Further, attendance-weighted mean-shift estimates shift upward within each class. As with ambulance arrivals, triangles lie to the right of circles across all classes, indicating that higher-volume sites see proportionally more high-acuity cases. The larger dot–triangle gaps in the rural sites indicate greater within-class heterogeneity among rural UEC sites, with small local sites appearing very different from larger rural sites.

High-acuity related distribution seen in Figure 4 may partially be due to the distribution of Type 1 EDs among the four classes. Supplementary Figure S1 presents annual acuity distributions for all departments and for Type 1 attendances alone. Within Type 1 departments, both rural classes had higher shares of high-acuity attendances (levels 1–3) than the urban classes in each financial year. For the Rural & Coastal class, this share increased from 49.8% in 2021/22 to 58.4% in 2023/24. These annual comparisons complement the pooled results in Figure 4 and show that class differences in recorded acuity remain visible when the analysis is restricted to major emergency departments.

The users in the most-deprived income quartile panel show that income deprivation is predominantly an urban phenomenon. We do not observe a difference between the “typical” site mean and the attendance-weighted site mean. The median age panel shows that the Rural & Coastal UEC sites serve the oldest population on average.

Taken together, the panels show a clear pattern between UEC classes. From the perspective of an average UEC site, urban sites handle more ambulance arrivals and higher-acuity attendances and serve more income-deprived populations, whereas rural sites serve older populations with lower acuity on average. However, when we weight the site average by site attendance volume, the difference between the two means indicates within class heterogeneity in UEC site attendances. The larger gaps between site and attendance-weighted means in the rural sites suggest additional pressure from ambulance arrivals and acuity, for the largest rural and coastal sites. Combined with the seasonal patterns observed earlier, this suggests that rural and coastal sites may experience distinct compound demand pressures across the year, both in terms of timing and case-mix.

## 4 Discussion

This study presents a transparent, utilisation-based approach for classifying emergency care sites based on the populations they serve, rather than by their postcode. Using patient residence at the small-area level, we constructed catchment-based site profiles and grouped sites into four classes: Rural & Coastal, Rural & Inland, Urban & Coastal, and Urban & Inland. This addresses an important practical gap for service planning and research.

Existing spatial classifications measure local population need, but fail to capture the operational context of UEC services. Provider postcode often diverges from catchment-area characteristics. For instance, an inland hospital may treat a substantial coastal cohort, whereas a coastal facility may draw heavily from inland urban centres. Classifying services solely by location thus misrepresents underlying demand, patient complexity, and seasonal volatility, distorting comparisons across providers and policy evaluations. Our catchment typology addresses this limitation, offering a transparent, reproducible framework to profile emergency services pressures at the UEC site level. Using the typology it is possible to compare demand, case-mix, and seasonal pressures in a way that is reproducible for policy, practice and research. It can also be used to support more informed decision-making on planning and resource allocation, with the potential to improve system performance and patient outcomes. This is particularly relevant for current national priorities to improve health in coastal communities and reduce geographical disparities, providing a mechanism through which progress against these objectives can be more transparently assessed (Chief Medical Officer for England, 2021).

The descriptive results show that UEC sites in Rural & Coastal, Rural & Inland, Urban & Coastal, and Urban & Inland differ not only in scale but also in the type of activity they perform. Urban UECs, especially Urban & Inland, have substantially higher attendance volumes and a larger share of ambulance arrivals and high-acuity attendances. Rural sites have lower average volume but show greater heterogeneity, with some larger rural UEC sites having a disproportionate share of arrivals by ambulance and high-acuity demand. This has important implications for how system pressures are presented and felt. Analyses based on the average site may suggest that rural UEC services face relatively lower acuity and demand intensity. However, from a system perspective, a large share of rural attendances is actually concentrated in a smaller number of higher-volume sites increasing their pressure. This difference is greater in rural and coastal UEC services, indicating that pressure is not evenly distributed across sites. In practice, this suggests that rural systems are not simply smaller versions of urban systems, they include a mixture of small local units and larger centres, and the latter may face similar pressure to urban major departments despite operating in more geographically dispersed settings.

Current approaches to managing UEC demand (ie. focus on winter pressures) may not fully capture the patterns observed in rural and coastal areas where demand increases more and most sharply during summer months and is concentrated in specific sites. As highlighted in recent national policy discussions on health in coastal communities, there is a need to move away from uniform, top-down approaches and towards models of care that reflect local population characteristics and service configurations (Chief Medical Officer for England, 2021). Our findings provide empirical support for this position and suggest that planning and resource allocation should account for both the uneven distribution of pressures across sites and the differing seasonal and case-mix profiles across geographic contexts.

The catchment-based typology and associated descriptive evidence provide a foundation for more targeted investigation of system performance and resilience. Our results reveal two distinct features of urgent and emergency care demand. First, baseline case mix and ambulance conveyance patterns differ systematically between urban and rural sites. Second, temporal demand surges concentrate heavily in Rural & Coastal UECs. From a policy perspective, the typology provides a practical mechanism to identify which UEC sites serve rural and coastal populations, addressing a current gap in national monitoring. This enables more consistent assessment of geographical disparities and supports evaluation of policy priority areas, including improving health in coastal communities and reducing inequalities in access and outcomes. By making service context observable at the provider level, it allows progress against these objectives to be more transparently assessed. From a service planning and design perspective, the findings suggest that a uniform approach to managing UEC demand may not be appropriate. Integrated care systems and providers can use the typology to distinguish between sites facing structurally different pressures, including differences in baseline case-mix and seasonal demand. This can inform more targeted resource allocation, capacity planning, and resilience strategies. For example, by identifying sites where summer demand surges coincide with high-acuity or ambulance arrivals, and where additional staffing, alternative care pathways, or service reconfiguration may be required. From a research perspective, the typology provides a consistent framework for analysing performance across heterogeneous service contexts, ready to be applied to other settings wherever an attendance-level dataset contains (i) a facility identifier and (ii) a small-area geography for users’ usual residence. It enables future work to examine metrics that directly capture system pressure, including waiting times, length of stay, admission rates, ambulance handover delays, and readmission patterns, and to test whether seasonal inflows translate into disproportionate crowding or delays in care in rural and coastal settings.

### 4.1 Limitations

This study is descriptive and relies on routine administrative data, so the typology and the patterns we report should be interpreted as characterising observed utilisation rather than underlying need. Catchments are inferred from attendances and may therefore be affected by access constraints, service availability, and patient choice. The approach does not directly capture unmet need or people who seek care outside the NHS emergency care system in England. Rurality and coastalness are assigned at LSOA level using standard lookups and a centroid-based coastal threshold, which can misclassify some locations near boundaries and does not account for travel routes or coastline accessibility. The three-year window may include short-term shocks or service changes, and seasonal patterns could differ in earlier or later periods.

## 5 Disclaimer

This paper uses anonymised patient-level data and linked geographic lookups to produce aggregated results. The interpretation and conclusions are those of the authors and do not necessarily reflect the views of the data providers or any affiliated organisations. The site typology presented here is a research construct intended to support analysis and monitoring and it should not be treated as an official designation of site or provider status.

## Data Availability

This work uses data provided by patients and collected by the NHS as part of their care and support. The Emergency Care Data Set (ECDS) is copyright 2017-2025, NHS England. Re-used with the permission of NHS England. All rights reserved.

## Acknowledgements and funding

This study is funded by the National Institute for Health and Care Research, Health and Social Care Delivery Research programme “Identifying innovative models of urgent and emergency care in rural and coastal areas in England” [NIHR162324].

The funder had no role in study design; collection, analysis and interpretation of data; writing of the report; or the decision to submit the article for publication.

The views expressed are those of the authors and not necessarily those of the NIHR.

This project was undertaken on the Data Safe Haven, which is an ISO 27001 certified environment for handling sensitive data, and is provided by the University of York. We are grateful for support by the York Data Safe Haven team and the Research Computing team.

Outputs are aggregate and are subject to disclosure control before release. No attempts are made to identify individuals or small groups. Ethical approval and data-sharing agreements cover the secondary analysis of ECDS and linkage to publicly available lookup files.

## Footnotes

1. Site mergers and acquisitions are available by request to the authors

2. Further details of the harmonisation procedures can be provided by request to the authors

## Appendix

### A ONS Rural–Urban Classification 2021

We use the Office for National Statistics (ONS) 2021 Rural–Urban Classification (RUC21) to characterise the rurality of users’ areas of residence at the LSOA21 level. RUC21 is a settlement-based statistical classification constructed from (i) residential address density and physical settlement form and (ii) relative access to major towns and cities. It is produced at Output Area (OA) level and then propagated to higher statistical geographies such as LSOAs and MSOAs.

RUC21 assigns each OA to one of three settlement types: Urban, Larger rural settlement, or Smaller rural settlement. OAs are classified as Urban if they have high residential address density or if they intersect with an Amalgamated Built Up Area (ABUA) of at least 10,000 residents; lower-density OAs (or those intersecting smaller ABUAs) are assigned to one of the two rural settlement categories. Each settlement type is further divided into a relative access class. “Nearer” indicates that an area can reach at least one Built Up Area (BUA) with population above 75,000 within 30 minutes travel by car (based on modelled travel time and the road network); “Further” indicates that it cannot. Table 2 shows the detailed breakdown of RUC21.

**Table 2:** Rural–Urban Classification 2021 (RUC21): settlement type by relative access.

| RUC21 category | Description |
| --- | --- |
| Urban, Nearer | Urban OA/LSOA, nearer to a major town/city |
| Urban, Further | Urban OA/LSOA, further from a major town/city |
| Larger rural settlement, Nearer | Larger rural settlement, nearer to a major town/city |
| Larger rural settlement, Further | Larger rural settlement, further from a major town/city |
| Smaller rural settlement, Nearer | Smaller rural settlement, nearer to a major town/city |
| Smaller rural settlement, Further | Smaller rural settlement, further from a major town/city |

For the main analyses we collapse RUC21 to a binary indicator, defining an LSOA21 as rural if it is classified as either a Larger rural settlement or Smaller rural settlement (Nearer or Further), and urban if it is classified as Urban (Nearer or Further). This aligns the rural/urban split with the settlement-based construction of RUC21.

### B Z-test

For each site *i* we compute the proportion of attendances whose users are rural (*p*^rural^_*i*_) and coastal (*p*^coast^_*i*_) over FY2021/22–FY2023/24. We compare each to the national mean (*p*^rural^_0_, = 0.146, *p*^coast^_0_ = 0.164) via one-sample z-tests for proportions:

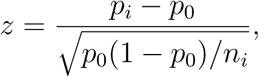

where *n_i_* is the site’s total attendances. Thresholds *z ≥* 1.96 and *z ≤ −*1.96 flag “meaningfully above” and “meaningfully below”. Combining rural and coastal flags yields four classes: *Rural & Coastal*, *Rural & Inland*, *Urban & Coastal*, *Urban & Inland*. “Buffer/mixed” sites (non-significant on one or both tests) are excluded from class-level summaries.

### C Department types of AED sites in ECDS

**Table 3:** Enhanced Technical Output Specification (ETOS): ECDS Department Types.

| Code | Description |
| --- | --- |
| 01 | Full resuscitation, designated accommodation, consultant led, 24 hours |
| 02 | Mono-specialty, designated accommodation, consultant led (eg. dental, ophthalmology) |
| 03 | Other type, A&E minor injury, designated accommodation, doctor/GP/nurse led, without appointment (eg. urgent treatment centres; excludes NHS walk-in centres) |
| 04 | NHS walk-in centres |
| 05 | Ambulatory emergency-care service (piloting only) |

